# Intrinsic and effective severity of COVID-19 cases infected with the ancestral strain and Omicron BA.2 variant in Hong Kong

**DOI:** 10.1101/2023.02.13.23285848

**Authors:** Jessica Y. Wong, Justin K. Cheung, Yun Lin, Helen S. Bond, Eric H. Y. Lau, Dennis K. M. Ip, Benjamin J. Cowling, Peng Wu

## Abstract

**Background:** Understanding severity of infections with SARS-CoV-2 and its variants is crucial to inform public health measures. Here we used COVID-19 patient data from Hong Kong to characterise the severity profile of COVID-19 and to examine factors associated with fatality of infection.

**Methods:** Time-varying and age-specific effective severity measured by case-hospitalization risk and hospitalization risk was estimated with all individual COVID-19 case data collected in Hong Kong from 23 January 2020 through to 26 October 2022 over six epidemic waves, in comparison with estimates of influenza A(H1N1)pdm09 during the 2009 pandemic. The intrinsic severity of Omicron BA.2 was compared with the estimate for the ancestral strain with the data from unvaccinated patients without previous infections. Factors potentially associated with the fatality risk of hospitalized Omicron patients were also examined.

**Results:** With 32,222 COVID-19 hospitalizations and 9,669 deaths confirmed over 6 epidemic waves in Hong Kong, the time-varying hospitalization fatality risk dramatically increased from below 10% before the largest fifth wave of Omicron BA.2, to 41% during the peak of the fifth wave when hospital resources were severely constrained. The age-specific fatality risk in unvaccinated hospitalized Omicron cases was comparable to the estimates for unvaccinated cases with the ancestral strain. During epidemics predominated by Omicron BA.2, the highest fatality risk was amongst unvaccinated patients aged ≥80 years and the risk was inversely associated with the number of vaccination doses received.

**Conclusions:** Omicron has comparable intrinsic severity to the ancestral Wuhan strain although the effective severity is substantially lower in Omicron cases due to vaccination. With a moderate-to-high coverage of vaccination, hospitalized COVID-19 patients caused by Omicron subvariants appeared to have similar age-specific risks of fatality to patients hospitalized with influenza A(H1N1)pdm09.

## INTRODUCTION

Hong Kong successfully suppressed four epidemic waves of infection with the ancestral strain of SARS-CoV-2 during the first two years of the pandemic, with 12,631 RT-PCR confirmed infections (1.7 cases per 1,000 population) and 213 deaths reported by the end of 2021 [1]. Serologic data confirmed that below 1% of the population had been infected over the same period [2]. However, a large community epidemic caused by Omicron subvariant peaked in early March 2022 [3, 4], leading to over 1.1 million confirmed cases (164.3 cases per 1,000 population) and 9,157 deaths. Two COVID-19 vaccines, the inactivated (CoronaVac, Sinovac) and the mRNA vaccine (BNT162b2, BioNTech/Fosun Pharma), were made available in Hong Kong in early 2021. As of 1 January 2022, approximately 70% of the Hong Kong population aged 3 years or above had received at least two doses of vaccination [5] although vaccination uptake was lower in older adults [6].

The severity profile of SARS-CoV-2 infections is one of the major determinants for assessing potential health impact of epidemics on a population [7]. One measure of infection severity is the “hospitalization fatality risk” (HFR). This describes the risk of death among hospitalized cases with laboratory-confirmed SARS-CoV-2 infections [8], which comprise a smaller and more homogenous subgroup of infected individuals requiring admission for healthcare than all laboratory-confirmed cases. A related measure of severity is the risk of hospitalization among all cases, i.e. the case hospitalization risk (CHR). The CHR multiplied by the HFR should approximate the fatality risk among all confirmed cases, i.e. the case fatality risk (CFR).

Here, we analyzed detailed individual case data from Hong Kong, to quantify the overall and temporal patterns of COVID-19 severity in Hong Kong, and identify factors associated with mortality among hospitalized COVID-19 cases. Because Omicron BA.2 was the first strain of COVID-19 to circulate widely in Hong Kong we also aim to estimate the intrinsic severity of SARS-CoV-2 variants using data on infections in unvaccinated individuals.

## METHODS

### Sources of data

Individual data on all laboratory-confirmed COVID-19 cases infected locally in Hong Kong from 23 January 2020 to 26 October 2022 were obtained from the Hospital Authority and Centre for Health Protection [5]. RT-PCR was used exclusively for confirmation of COVID-19 until 25 February 2022 when a positive rapid antigen test was also accepted as laboratory confirmation [9]. From 7 June 2022 onwards, individuals reporting a positive rapid antigen test were issued with a compulsory testing notice and required to undergo PCR testing within the next two days, and any individuals with negative PCR tests were not counted as COVID-19 cases [10]. Demographic, clinical and epidemiological information was collected for individual patients, including age, sex, comorbidities, oxygen saturation levels measured by the pulse oximetry in hospital, severity outcome at discharge, vaccination status, use of nirmatrelvir-ritonavir (Paxlovid) or molnupiravir, residence in care homes for the elderly (RCHE), dates of COVID-19 vaccination for individual doses, laboratory confirmation, hospital admission, death and discharge. Our study received ethical approval from the Institutional Review Board of the University of Hong Kong. Up to 14 February 2022, all laboratory-confirmed COVID-19 cases in Hong Kong were admitted into hospital for isolation, and from 15 February onwards, given the rapidly increasing number of COVID-19 cases and limited hospital capacity, only those requiring hospital medical care were admitted. In this study, we defined hospitalized cases as COVID-19 patients confirmed by RT-PCR or rapid antigen test and clinically classified as a severe, critically ill or fatal case based on assessment of oxygen desaturation, use of medication and procedure, and information such as A&E departmental visits and ICU admissions from clinical records, i.e., cases requiring healthcare in hospital. Detailed information on severity classification of COVID-19 cases is shown in Appendix. We used data on patients with laboratory confirmation of SARS-CoV-2 infection obtained in the 14 days preceding hospital admission in the analysis on HFR, and used data on all patients tested positive for SARS-CoV-2 either by RT-PCR or rapid antigen test in the analysis of CHR.

For comparison, we used the data obtained from the Hospital Authority on all individual patients with laboratory-confirmed A(H1N1)pdm09 infection identified during May-December 2009 when influenza A(H1N1)pdm09 virus was first circulating and monovalent H1N1pdm09 vaccines had not been widely used in Hong Kong [11], including age, sex, dates of admission into hospital and intensive care unit, dates of receipt of oseltamivir treatment and death.

### Statistical analysis

All COVID-19 cases confirmed in each epidemic wave or period of wave were examined in terms of age, sex, severity status and delay distributions. We estimated the effective severity of COVID-19 with the case hospitalization risk (CHR) and the hospitalization fatality risk (HFR). The weekly CHR was estimated as the reported weekly number of hospitalized COVID-19 cases divided by the weekly total number of confirmed cases allowing for a mean lag of 3 days from case notification to hospitalization. The weekly HFR was calculated as the proportion of fatal cases (eventual number of deaths occurred by the end of the study period) among all hospitalized cases admitted in a particular week. We estimated the CHR and HFR in time periods during which at least 35 confirmed cases and at least 15 confirmed hospitalized cases were reported per week in 4 or more consecutive weeks in order to provide effective comparisons between different epidemic waves avoiding large uncertainties from very small case numbers. We also estimated the age-stratified CHR and HFR with the same data for five age groups: <18 years, 18-44 years, 45-64 years, 65-79 years and ≥80 years for each pre-defined epidemic wave over the study period.

The data on COVID-19 cases without vaccination in Hong Kong before widespread of Omicron in wave 5 particularly in older adults with a low vaccination coverage allowed us to assess the intrinsic severity of SARS-CoV-2. We determined the HFR among hospitalized cases at age of <65, 65-79 and ≥80 years with complete comorbidity information. We also estimated the age-specific relative risk in logarithmic scale as the HFR in the early part of the fifth wave divided by the HFR in waves 1-4. Cases included in the analysis were confirmed before 14 February 2022 without receiving any dose of COVID-19 vaccination more than 14 days prior to confirmation of COVID-19 and without a recorded previous infection. We applied logistic regression models to the daily unvaccinated hospitalized COVID-19 cases confirmed in the early part of the fifth wave and earlier epidemic waves to examine factors potentially associated with the intrinsic fatality risk of SARS-CoV-2. In the regression model, we adjusted for age group, gender, comorbidities, and RCHE status. Age-specific HFR estimates were also obtained for unvaccinated cases confirmed in wave 6 for comparison.

Omicron BA.2 caused the largest epidemic wave 5 in Hong Kong since January 2022 extending to wave 6 in May-August 2022. We estimated the HFR with cases confirmed during the period predominated by Omicron BA.2 by vaccine dose and type to illustrate the potential protection provided by COVID-19 vaccination. We determined the vaccine-related relative risk in logarithmic scale for the three age groups as the HFR among cases with a specific vaccine dose combination divided by the HFR among cases that did not receive any dose of COVID-19 vaccination. We performed a similar regression analysis to compare the risk of mortality for patients confirmed in wave 5. In the model, we further adjusted for vaccination type and dose, antiviral use and period of case confirmation.

We estimated the HFR of influenza A(H1N1)pdm09 with cases confirmed in 2009 as the division of cumulative number of deaths by the cumulative number of laboratory-confirmed hospitalized cases who were treated with oseltamivir or admitted into an intensive care unit during hospital stay. All analyses were conducted with R version 4.1.0 (R Foundation for Statistical Computing, Austria).

## RESULTS

The periods of spread of SARS-CoV-2 in Hong Kong can be divided into six epidemic waves, including four prior to the widespread uptake of vaccines, a large fifth wave predominated by Omicron BA.2 between January and May 2022, and a sixth wave driven by Omicron BA.2 and BA.4/5 subvariants (Figure 1). A total of 1,979, 22,805 and 7,438 COVID-19 cases were hospitalized for treatment in waves 1-4, wave 5 and wave 6, respectively. Of all the deaths, 88.8% (8,586/9,669) occurred within 28 days of confirmation, and 98.7% (9,548/9,669) occurred within 90 days. In the peak period of wave 5 (wave 5b, between 14 February and 29 April 2022), 0.8% (4/491), 9.6% (62/646), 22.7% (568/2,504), 31.5% (1,887/5,986) and 46.0% (5,832/12,667) of the hospitalized cases were fatal in age groups <18, 18-44, 45-64, 65-79 and ≥80 years, respectively (Appendix). The median admission-to-death intervals decreased from 18 days in waves 1-4 to 8 days in the peak period of wave 5. The median admission-to-discharge intervals were very similar, decreasing from 17 days to 11 days in waves 1-4 to peak period of wave 5 respectively.

**Figure 1:**
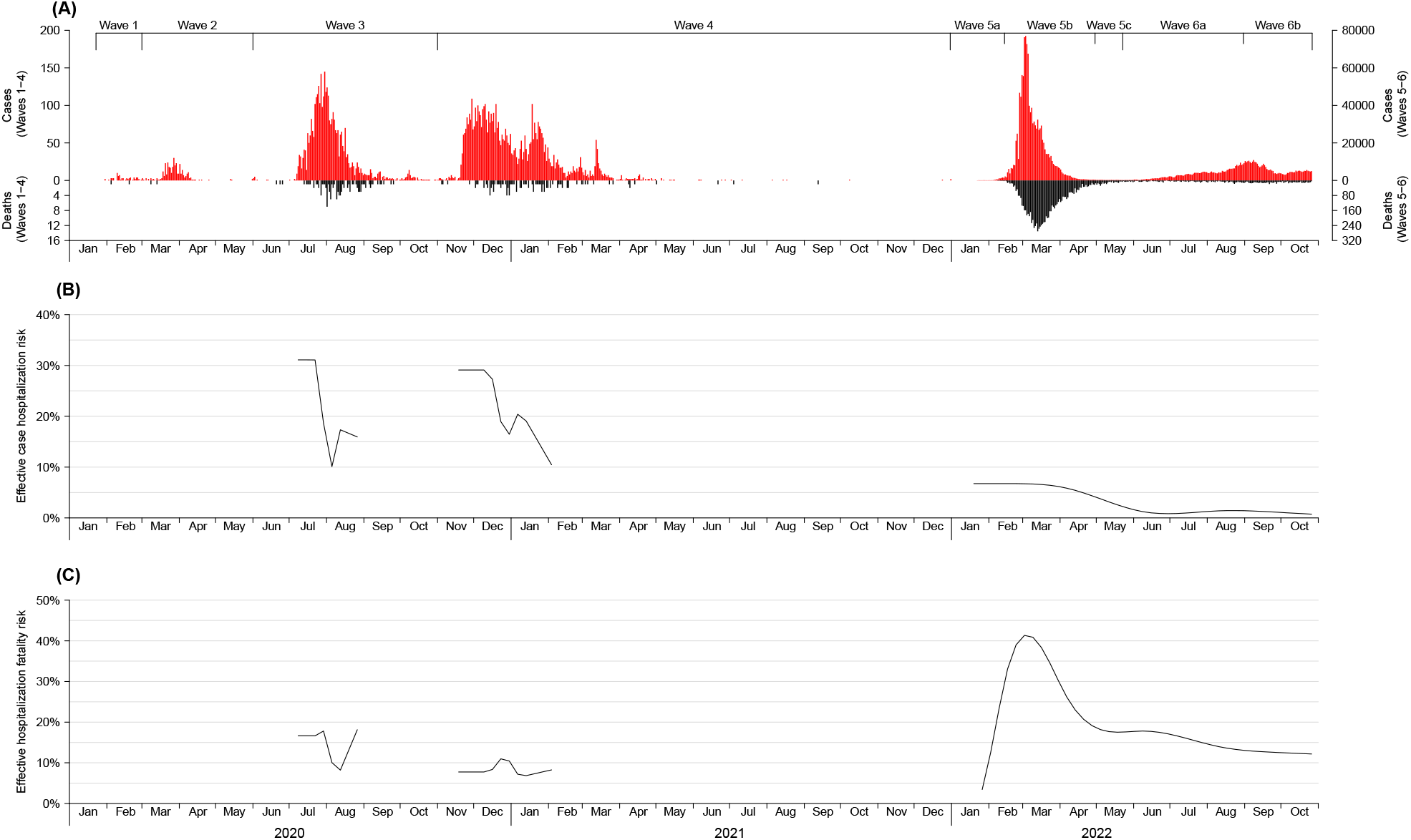
Local COVID-19 cases and deaths, estimated case hospitalization risk and estimated hospitalization fatality risk in Hong Kong, 2020-2022. (A) Epidemic curve of daily local COVID-19 cases and deaths in wave 1 to wave 6. Cases were stratified by date of confirmation and deaths were stratified by date of death. (B) Weekly effective case hospitalization risk. (C) Weekly effective hospitalization fatality risk. Estimates in panels (B) and (C) were only plotted for weeks when ≥35 local cases and ≥15 hospitalized cases were confirmed for at least 4 consecutive weeks.

From January 2020 to October 2022, the smoothed estimate of weekly CHR gradually decreased from 15.9% at the end of wave 3 to 10.5% at the end of wave 4 (Figure 1). In comparison, the weekly HFR estimates similarly declined from 18.1% (95% CI: 9.2%, 27.1%) to 8.3% (95% CI: 3.0%, 13.5%) during the same time period. In waves 5-6, CHR estimates were relatively stable at low levels (around 0.7%) while the HFR estimates increased substantially from 3.4% (95% CI: 0.0%, 7.7%) in late January 2022, peaked at 41.3% (95% CI: 37.9%, 44.8%) in early March before declining to 12.2% (95% CI: 7.9%, 16.4%) by the end of October.

The highest hospitalization risk was observed in cases aged ≥65 years (8.2%, 95% CI: 8.1%, 8.3%) in wave 5, similar to cases at the same age (5.2%, 95% CI: 5.0%, 5.3%) in wave 6 (Figure 2). The CHR of cases aged 45-64 and ≥65 years in waves 1-4 appeared to be approximately six times the risk in those at the same age in wave 5 (50.0%, 95% CI: 47.8%, 52.4%). The fatality risk of hospitalized cases aged ≥65 years (41.1%, 95% CI: 40.4%, 41.8%) in wave 5 doubled the risk in individuals in the same age group in waves 1-4 (19.7%, 95% CI: 17.2%, 22.4%) and wave 6 (17.0%, 95% CI: 16.0%, 18.0%), and the pattern remained the same for 45-64 years. The HFR estimates were comparable between hospitalized influenza A(H1N1)pdm09 cases in 2009 and COVID-19 cases confirmed during wave 6 (Figure 2B).

**Figure 2:**
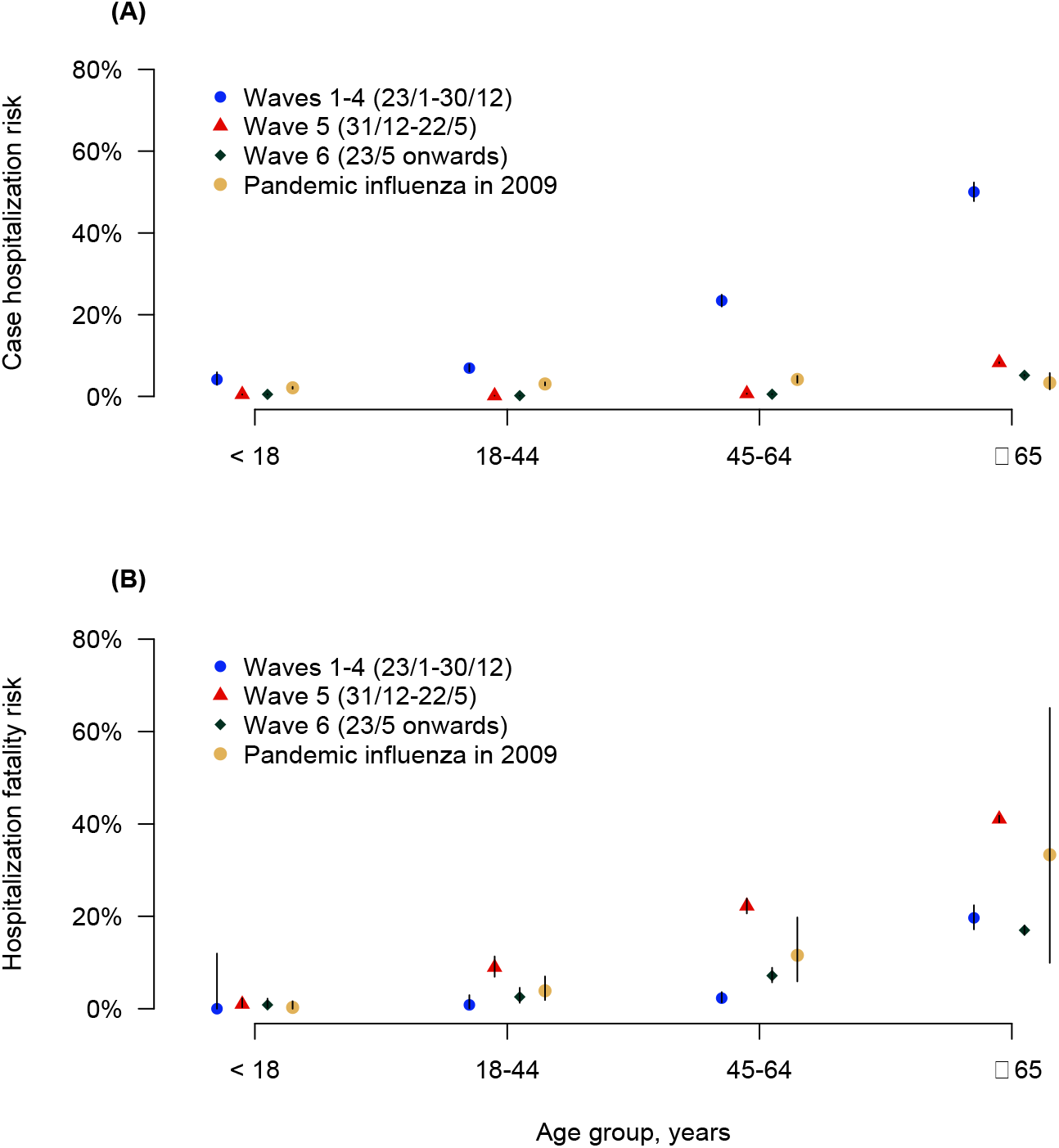
Age-specific estimates of the case hospitalization and hospitalization fatality risks for COVID-19 in waves 1-4, wave 5 and wave 6 in Hong Kong, compared with estimates of the hospitalization fatality risk for all hospitalized patients with pandemic influenza A(H1N1)pdm09 in 2009. (A) Case hospitalization risk in waves 1-4, 5 and 6 by age group, compared with pandemic influenza (H1N1)pdm09 in 2009. (B) Hospitalization fatality risk in waves 1-4, 5 and 6 by age group, compared with pandemic influenza (H1N1)pdm09 in 2009.

There were 22,604 COVID-19 patients hospitalized in wave 5 between 31 December 2021 and 22 May 2022 (Table 1). After accounting for factors associated with mortality, individuals with at least one dose of BNT162b2 or CoronaVac had significantly lower odds of mortality than those unvaccinated, with the lowest odds observed in individuals with three doses of BNT162b2 (OR: 0.31, 95% CI: 0.18, 0.49). In addition to at an older age (65-79 years and ≥80 years vs 45-64 years), being an RCHE resident, having malignant neoplasms or cardiovascular diseases, and confirmed during the peak period were associated with an increased odds of mortality. Female cases, at a younger age or being treated with antivirals (Paxlovid and molnupiravir) had a lower odds of being fatal.

**Table 1.**
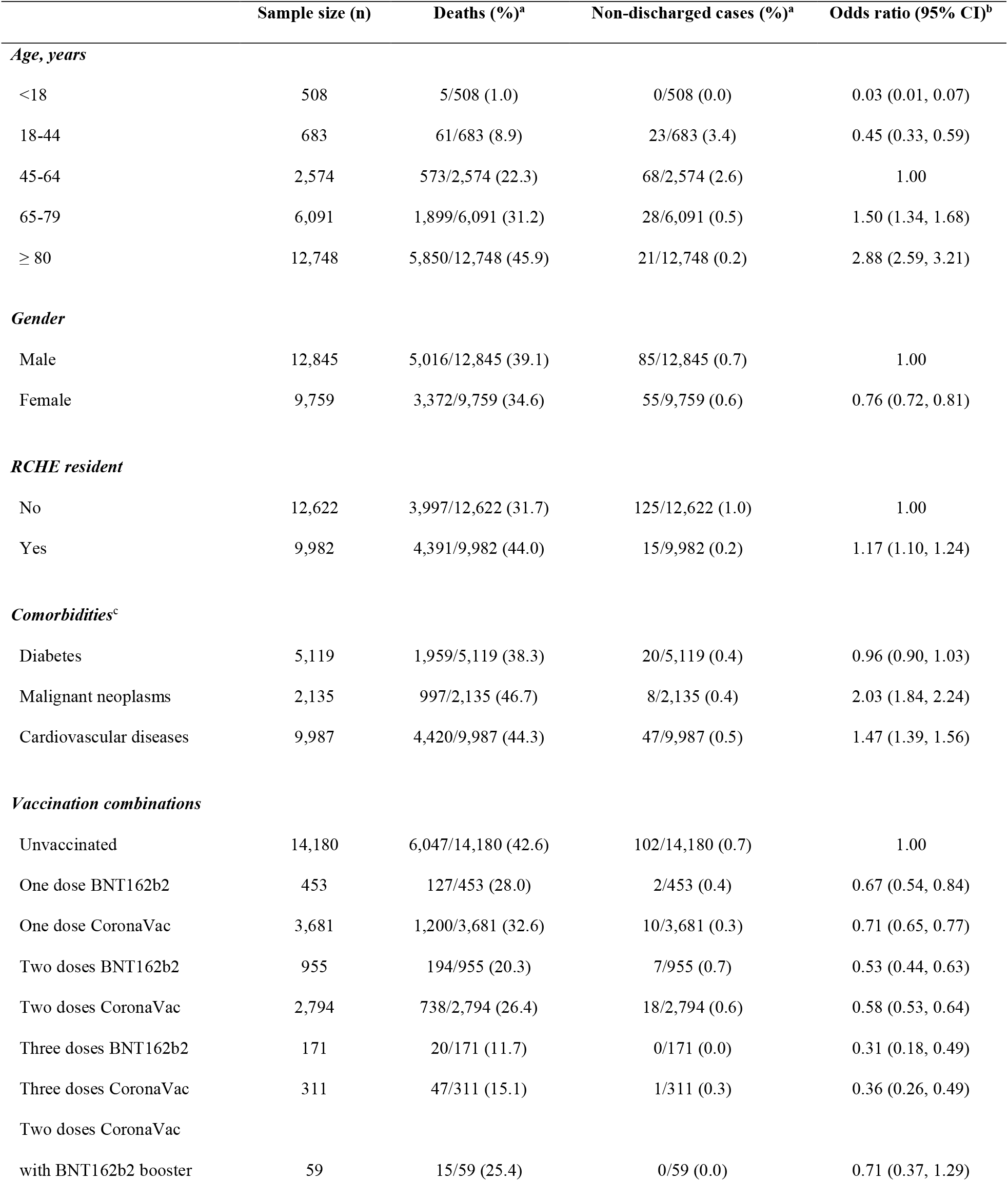
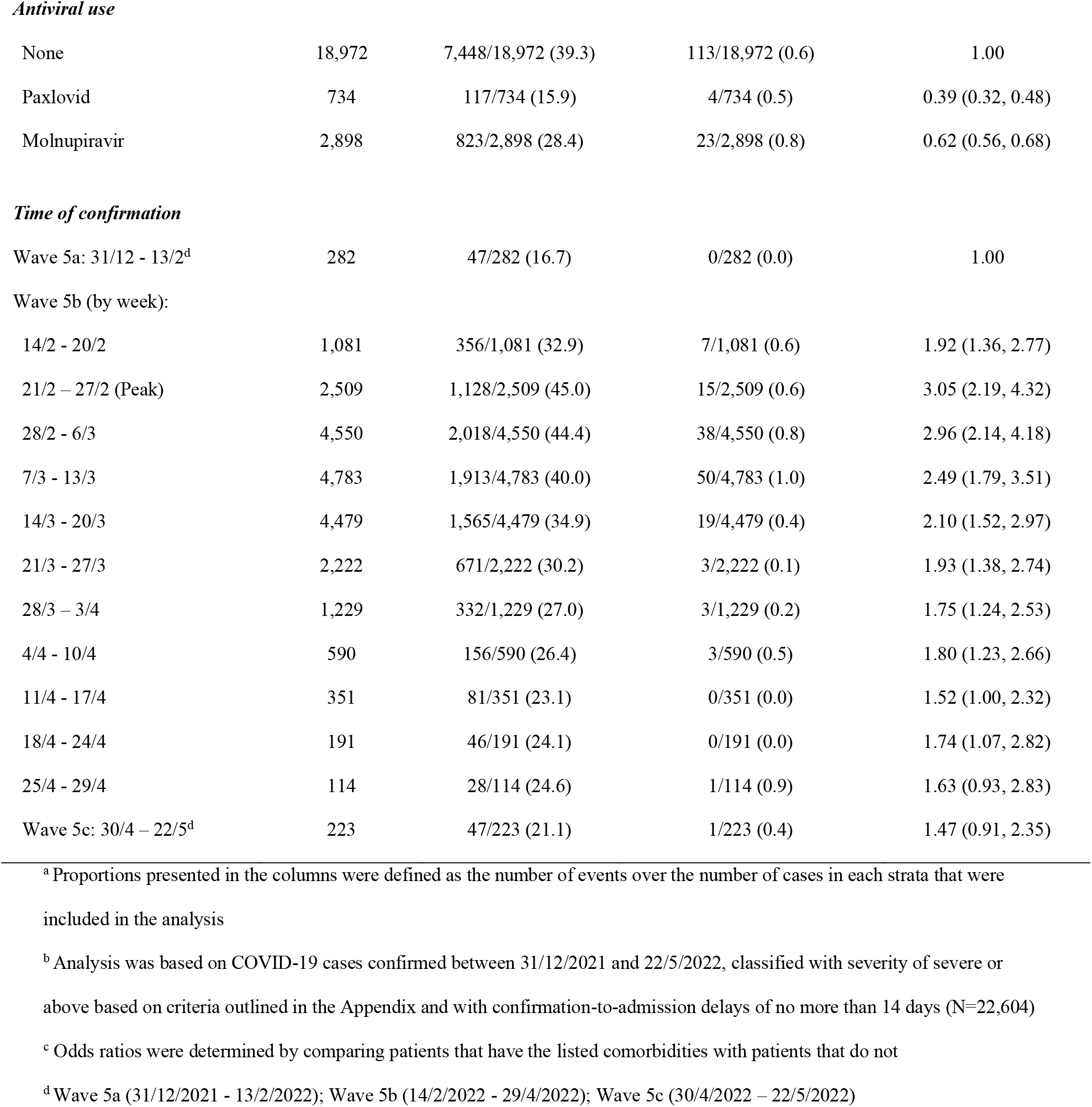
Logistic regression of risk factors of mortality among hospitalized local COVID-19 cases confirmed in wave 5 predominated by Omicron subvariants from 31 December 2021 to 22 May 2022.

|  | Sample size (n) | Deaths (%)<sup>a</sup> | Non-discharged cases (%)<sup>a</sup> | Odds ratio (95% CI)<sup>b</sup> |
| --- | --- | --- | --- | --- |
| ***Age, years*** |  |  |  |  |
| <18 | 508 | 5/508 (1.0) | 0/508 (0.0) | 0.03 (0.01, 0.07) |
| 18-44 | 683 | 61/683 (8.9) | 23/683 (3.4) | 0.45 (0.33, 0.59) |
| 45-64 | 2,574 | 573/2,574 (22.3) | 68/2,574 (2.6) | 1.00 |
| 65-79 | 6,091 | 1,899/6,091 (31.2) | 28/6,091 (0.5) | 1.50 (1.34, 1.68) |
| ≥ 80 | 12,748 | 5,850/12,748 (45.9) | 21/12,748 (0.2) | 2.88 (2.59, 3.21) |
| ***Gender*** |  |  |  |  |
| Male | 12,845 | 5,016/12,845 (39.1) | 85/12,845 (0.7) | 1.00 |
| Female | 9,759 | 3,372/9,759 (34.6) | 55/9,759 (0.6) | 0.76 (0.72, 0.81) |
| ***RCHE resident*** |  |  |  |  |
| No | 12,622 | 3,997/12,622 (31.7) | 125/12,622 (1.0) | 1.00 |
| Yes | 9,982 | 4,391/9,982 (44.0) | 15/9,982 (0.2) | 1.17 (1.10, 1.24) |
| ***Comorbidities<sup>c</sup>*** |  |  |  |  |
| Diabetes | 5,119 | 1,959/5,119 (38.3) | 20/5,119 (0.4) | 0.96 (0.90, 1.03) |
| Malignant neoplasms | 2,135 | 997/2,135 (46.7) | 8/2,135 (0.4) | 2.03 (1.84, 2.24) |
| Cardiovascular diseases | 9,987 | 4,420/9,987 (44.3) | 47/9,987 (0.5) | 1.47 (1.39, 1.56) |
| ***Vaccination combinations*** |  |  |  |  |
| Unvaccinated | 14,180 | 6,047/14,180 (42.6) | 102/14,180 (0.7) | 1.00 |
| One dose BNT162b2 | 453 | 127/453 (28.0) | 2/453 (0.4) | 0.67 (0.54, 0.84) |
| One dose CoronaVac | 3,681 | 1,200/3,681 (32.6) | 10/3,681 (0.3) | 0.71 (0.65, 0.77) |
| Two doses BNT162b2 | 955 | 194/955 (20.3) | 7/955 (0.7) | 0.53 (0.44, 0.63) |
| Two doses CoronaVac | 2,794 | 738/2,794 (26.4) | 18/2,794 (0.6) | 0.58 (0.53, 0.64) |
| Three doses BNT162b2 | 171 | 20/171 (11.7) | 0/171 (0.0) | 0.31 (0.18, 0.49) |
| Three doses CoronaVac | 311 | 47/311 (15.1) | 1/311 (0.3) | 0.36 (0.26, 0.49) |
| Two doses CoronaVac<br>with BNT162b2 booster | 59 | 15/59 (25.4) | 0/59 (0.0) | 0.71 (0.37, 1.29) |
| **Antiviral use** |  |  |  |  |
| None | 18,972 | 7,448/18,972 (39.3) | 113/18,972 (0.6) | 1.00 |
| Paxlovid | 734 | 117/734 (15.9) | 4/734 (0.5) | 0.39 (0.32, 0.48) |
| Molnupiravir | 2,898 | 823/2,898 (28.4) | 23/2,898 (0.8) | 0.62 (0.56, 0.68) |
| **Time of confirmation** |  |  |  |  |
| Wave 5a: 31/12 - 13/2<sup>d</sup> | 282 | 47/282 (16.7) | 0/282 (0.0) | 1.00 |
| Wave 5b (by week): |  |  |  |  |
| 14/2 - 20/2 | 1,081 | 356/1,081 (32.9) | 7/1,081 (0.6) | 1.92 (1.36, 2.77) |
| 21/2 – 27/2 (Peak) | 2,509 | 1,128/2,509 (45.0) | 15/2,509 (0.6) | 3.05 (2.19, 4.32) |
| 28/2 - 6/3 | 4,550 | 2,018/4,550 (44.4) | 38/4,550 (0.8) | 2.96 (2.14, 4.18) |
| 7/3 - 13/3 | 4,783 | 1,913/4,783 (40.0) | 50/4,783 (1.0) | 2.49 (1.79, 3.51) |
| 14/3 - 20/3 | 4,479 | 1,565/4,479 (34.9) | 19/4,479 (0.4) | 2.10 (1.52, 2.97) |
| 21/3 - 27/3 | 2,222 | 671/2,222 (30.2) | 3/2,222 (0.1) | 1.93 (1.38, 2.74) |
| 28/3 – 3/4 | 1,229 | 332/1,229 (27.0) | 3/1,229 (0.2) | 1.75 (1.24, 2.53) |
| 4/4 - 10/4 | 590 | 156/590 (26.4) | 3/590 (0.5) | 1.80 (1.23, 2.66) |
| 11/4 - 17/4 | 351 | 81/351 (23.1) | 0/351 (0.0) | 1.52 (1.00, 2.32) |
| 18/4 - 24/4 | 191 | 46/191 (24.1) | 0/191 (0.0) | 1.74 (1.07, 2.82) |
| 25/4 - 29/4 | 114 | 28/114 (24.6) | 1/114 (0.9) | 1.63 (0.93, 2.83) |
| Wave 5c: 30/4 – 22/5<sup>d</sup> | 223 | 47/223 (21.1) | 1/223 (0.4) | 1.47 (0.91, 2.35) |

<sup>a</sup> Proportions presented in the columns were defined as the number of events over the number of cases in each strata that were included in the analysis
<sup>b</sup> Analysis was based on COVID-19 cases confirmed between 31/12/2021 and 22/5/2022, classified with severity of severe or above based on criteria outlined in the Appendix and with confirmation-to-admission delays of no more than 14 days (N=22,604)
<sup>c</sup> Odds ratios were determined by comparing patients that have the listed comorbidities with patients that do not
<sup>d</sup> Wave 5a (31/12/2021 - 13/2/2022); Wave 5b (14/2/2022 - 29/4/2022); Wave 5c (30/4/2022 – 22/5/2022)

There were 1,974 and 186 unvaccinated COVID-19 patients hospitalized in waves 1-4 and wave 5a, respectively, included in our analysis of intrinsic severity (Table 2). The age-specific estimates of HFR for cases ≥80 years (39.7%, 95% CI: 28.5%, 51.9%) and 65-79 years (19.3%, 95% CI: 10.0%, 31.9%) confirmed in wave 5a were similar to the corresponding estimates of 39.0% (95% CI: 33.1%, 45.0%) and 11.4% (95% CI: 9.0%, 14.1%) for cases in waves 1-4 (Figure 3). No significant differences in fatality risk were observed for hospitalized cases in wave 5a relative to those in waves 1-4 across all age groups. After accounting for factors potentially associated with mortality, hospitalized cases confirmed in wave 5a showed a similar fatality risk to the cases from waves 1-4 (OR: 1.08, 95% CI: 0.61, 1.83). The odds of mortality were significantly higher in individuals aged 65-79 years (OR: 4.80, 95% CI: 2.94, 8.26) and ≥80 years (OR: 20.81, 95% CI: 12.66, 35.95) than in individuals aged 45-64 years.

**Table 2.** Logistic regression of risk factors of mortality among hospitalized local COVID-19 cases that did not receive any dose of COVID-19 vaccine, waves 1-4 vs wave 5a.

|  | Sample size (n) | Deaths (%)<sup>a</sup> | Non-discharged cases (%)<sup>a</sup> | Odds ratio (95% CI)<sup>b</sup> |
| --- | --- | --- | --- | --- |
| Age, years |  |  |  |  |
| <18 | 45 | 0/45 (0.0) | 0/45 (0.0) | 0.00 (0.00, 0.00) |
| 18-44 | 256 | 2/256 (0.8) | 0/256 (0.0) | 0.41 (0.06, 1.43) |
| 45-64 | 806 | 19/806 (2.4) | 1/806 (0.1) | 1.00 |
| 65-79 | 708 | 85/708 (12.0) | 0/708 (0.0) | 4.80 (2.94, 8.26) |
| ≥ 80 | 345 | 135/345 (39.1) | 1/345 (0.3) | 20.81 (12.66, 35.95) |
| Gender |  |  |  |  |
| Male | 1,222 | 146/1,222 (11.9) | 2/1,222 (0.2) | 1.00 |
| Female | 938 | 95/938 (10.1) | 0/938 (0.0) | 0.68 (0.49, 0.92) |
| RCHE resident |  |  |  |  |
| No | 2,107 | 220/2,107 (10.4) | 2/2,107 (0.1) | 1.00 |
| Yes | 53 | 21/53 (39.6) | 0/53 (0.0) | 1.39 (0.63, 3.06) |
| Comorbidities |  |  |  |  |
| None | 1,493 | 85/1,493 (5.7) | 2/1,493 (0.1) | 1.00 |
| At least one comorbidity<sup>c</sup> | 667 | 156/667 (23.4) | 0/667 (0.0) | 2.89 (2.13, 3.95) |
| Confirmation period<sup>d</sup> |  |  |  |  |
| Waves 1-4 | 1,974 | 198/1,974 (10.0) | 2/1,974 (0.1) | 1.00 |
| Wave 5a | 186 | 43/186 (23.1) | 0/186 (0.0) | 1.08 (0.61, 1.83) |

<sup>a</sup> Proportions presented in the columns were defined as the number of events over the number of cases in each strata that were included in the analysis
<sup>b</sup> Analysis was based on COVID-19 cases with dates of confirmation during Waves 1-4 or Wave 5a, classified with severity of severe or above based on criteria outlined in the Appendix and with confirmation-to-admission delays of no more than 14 days (N=2,160)
<sup>c</sup> If patients have at least one of the following comorbidities: diabetes, malignant neoplasms and cardiovascular diseases
<sup>d</sup> Waves 1-4 (23/1/2020 – 30/12/2021); Wave 5a (31/12/2021 – 13/2/2022)

**Figure 3:**
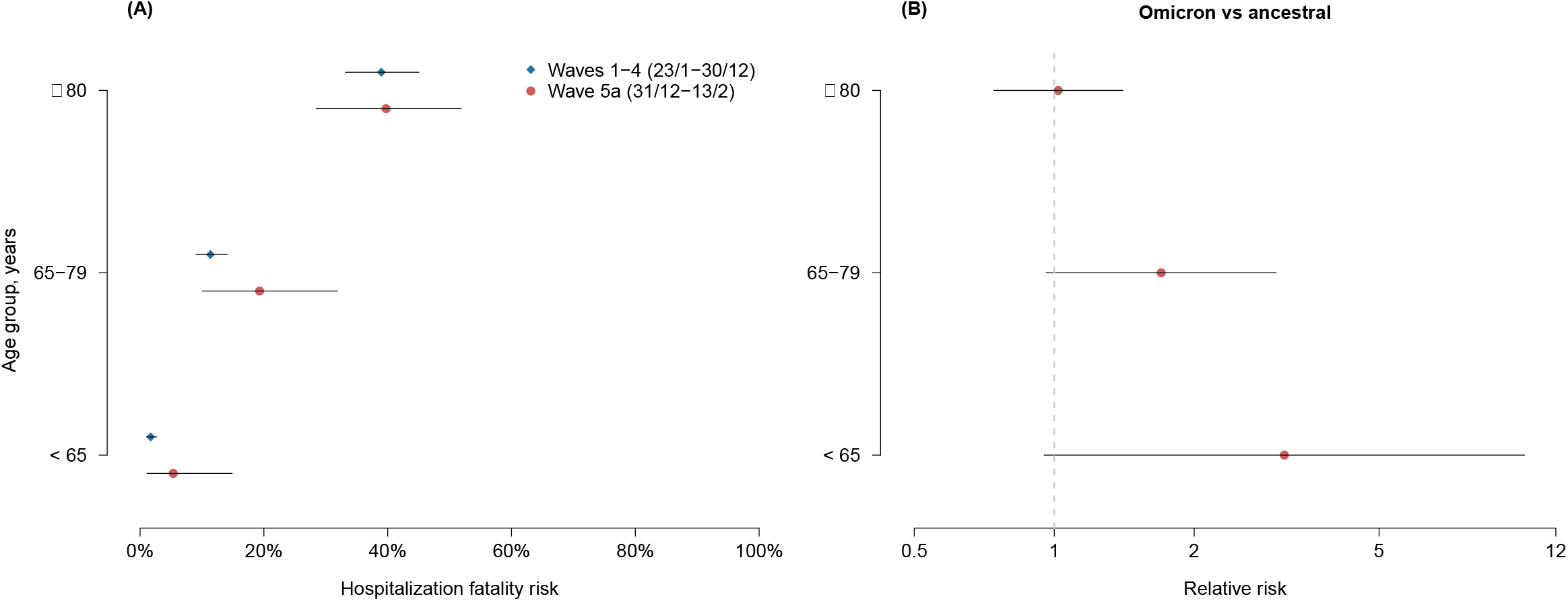
Age-specific estimates of the hospitalization fatality risk for COVID-19 among unvaccinated patients in Hong Kong by waves and relative risks of hospitalization fatality risk in wave 5a compared to waves 1-4. (A) Fatality risk among unvaccinated hospitalized local COVID-19 cases, in waves 1-4 compared with wave 5a by age group. (B) Relative risk of hospitalization fatality risk in wave 5a compared to waves 1-4 by age group.

Among cases hospitalized in non-peak periods of wave 5 and the first half of wave 6 (wave 5a from 31 December 2021 to 13 February 2022, and waves 5c and 6a from 30 April to 30 August 2022) with Omicron BA.2 predominant, mean estimates of the fatality risk were largely higher in unvaccinated patients and those receiving fewer doses of vaccine in age groups <65, 65-79 and ≥80 years except for those with two doses of BNT162b2 showing a higher mean estimate of HFR than cases receiving one dose only or 2-dose CoronaVac among cases aged <65 years (Appendix). Relative risk estimates showed that vaccination had a considerable effect in reducing fatality risk, especially for those who received at least two doses of CoronaVac or BNT162b2 in ≥80 years.

## DISCUSSION

Infections with the Omicron variant were originally recognized as being milder than with other variants [12] although it has been challenging to determine whether the attenuated severity of Omicron infections is due to an increased competency in infecting and replicating in the upper respiratory tract [13], or acquired immunity from previous infection and/or vaccination in affected populations [14]. Our analysis of intrinsic severity was conducted based on fatalities in unvaccinated COVID-19 patients admitted into hospital for treatment in Hong Kong during epidemic waves predominated by the ancestral Wuhan strain or Omicron BA.2 during January 2020 – February 2022 with a relatively low population coverage of COVID-19 vaccines especially in the elderly. The estimates of HFR indicated that Omicron BA.2 had a similar intrinsic severity to infections with the ancestral strain (Table 2).

Comparatively lower fatality risk in hospitalized COVID-19 patients infected with the ancestral strain or Omicron than Delta were reported in unvaccinated individuals from the UK and the US [15-18].

In estimation of fatality risk of COVID-19, using HFR was perhaps less biased than CFR using the number of confirmed cases as the denominator because the ascertainment of cases in need of hospital care would be relatively homogenous particularly in places with adequate capacity in case identification and healthcare services. Non-pharmaceutical and pharmaceutical measures have been widely used to slow down the transmission of COVID-19 to “flatten the curve” particularly in order to reduce the peak demand for hospital services. However, when healthcare services are under severe pressure often due to rapidly spread of infection, both hospitalized patient numbers and outcomes could be heavily affected, which would impact the HFR. Extreme pressure on hospital resources during large surges in COVID hospitalizations has been estimated to cause significantly increased COVID mortality rates in some locations, including Italy, the United States, England, and Brazil [19-23]. We estimated that the estimated HFR increased by a factor of three during the peak of the Omicron BA.2 wave in Hong Kong (Table 1). Therefore, in our analysis, we defined hospitalized COVID-19 patients with a set of pre-determined criteria (Appendix) to allow for comparison of HFR estimates across epidemic waves.

It is well established that the severity of COVID-19 increases substantially with age [24] as also shown in our age-specific estimates of HFR. The age-specific HFRs in our study indicated that a much higher fatality risk in adult cases hospitalized in the epidemic wave of Omicron than earlier waves, different from previous studies showing reduction in disease severity [25, 26]. This likely resulted from a higher proportion of older adults infected in wave 5 (Appendix) within a short time period leading to less optimal clinical outcomes in patients with constrained healthcare capacity [27].

By the end of wave 4 in December 2021, only around 12,000 COVID-19 cases were confirmed in Hong Kong [28], with a low vaccine coverage with two or more doses of either CoronaVac or BNT162b2 particularly in those ≥80 years of age [5]. We had a unique opportunity to investigate the intrinsic HFR for the Omicron in comparison with the ancestral strain by making use of fatality information collected from unvaccinated individuals, and at the same time to explore the potential protection against death from vaccines using vaccinated COVID-19 cases without a recorded infection history. Our estimation overcame some challenges in understanding the intrinsic severity of Omicron [14] while further virological and immunological evidence is needed to back up the observations from epidemiologic studies. However, we were only able to estimate intrinsic severity of Omicron BA.2 here because COVID-19 had not circulated widely in Hong Kong prior to 2022.

The estimated lower risk of death among hospitalized COVID-19 cases was indicated for vaccinees with any dose of the vaccine in comparison to the unvaccinated inpatients (Table 1), largely comparable with our previous findings on the effectiveness of COVID-19 vaccines against severe COVID-19 [5]. Furthermore, the potential protection provided by Paxlovid or molnupiravir demonstrated in a local territory-wide cohort of COVID-19 inpatients and outpatients during the Omicron BA.2 wave [29, 30] perhaps could explain the lower HFR estimated in wave 6 compared with wave 5c (Appendix) considering a gradually expanding use of the antivirals since mid-March 2022 in Hong Kong [31].

There are several limitations in our study. First, temporal comparison of severity estimates over epidemic waves might be affected by varied case definitions and practices in hospital admission over time [32]. We classified the severity status of laboratory-confirmed COVID-19 cases based on objective measures as much as possible, and defined hospitalized cases consistently throughout the study period, aiming to minimize bias in the denominator of HFR, and enable valid comparisons of severity measures across waves and patient groups. Second, fatality risk might differ in patients admitted due to COVID-19 and those hospitalized for other reasons and infected later with COVID-19, which might affect estimates of the HFR [25]. We limited our analysis on HFR to hospitalized patients with the confirmation 14 days earlier than the admission to exclude patients who might be infected in hospitals.

Third, clinical outcomes were unknown for 1.6% of the patients who were still in hospital by the end of the study. The exact estimates of HFR for wave 6b might change slightly if final outcomes of these patients are obtained although we do not anticipate this would change our study conclusions given the small number of patients involved.

In conclusion, similar intrinsic severity of Omicron to the ancestral strain and the protection conferred by vaccines against fatality risk in hospitalized COVID-19 patients highlighted that vaccination is critically important in reducing COVID-19 associated health impact. Continued monitoring of epidemiologic characteristics of new variants and subvariants of SARS-CoV-2 is crucial, including assessment of effective severity. As testing and reporting cases in the community reduces in frequency, but hospital testing likely continues in many locations, data on the HFR may provide a metric that is easier to evaluate over time than CFR. Serologic data on patterns in infections over time, if available, could provide a more complete picture of changes over time in severity.

## Data Availability

Restrictions apply to the availability of these data. The hospitalization and mortality data are available for access with the permission from the Hospital Authority and the Census and Statistics Department of Hong Kong.

## ACKNOWLEDGMENTS

The authors thank Julie Au for technical support.

## AUTHOR CONTRIBUTIONS

The study was conceived by BJC and PW. JYW, PW and JKC analyzed the data. JYW wrote the first draft of the manuscript. All authors provided critical review and revision of the text and approved the final version.

