## Appendix for "Intrinsic and effective severity of COVID-19 cases infected with the ancestral strain and Omicron BA.2 variant in Hong Kong"

This appendix provides additional information on circulating variants in Hong Kong, case identification practices of confirmed cases throughout the pandemic and data used in the study.

### 1. Circulating variants in Hong Kong

Hong Kong has successfully controlled the first four waves of the pandemic through a combination of non-pharmaceutical measures. Prior to 31 December 2021, two wild-type lineages B.1.1.63 and B.1.36.27 predominated the pandemic in July and November 2020 respectively and circulated thereafter. Variants of concern were not detected among local COVID-19 cases throughout 2020 and 2021 [1, 2]. Low levels of immunity at the population level contributed to a massive fifth primary Omicron wave predominated by the BA.2 variant, followed by a secondary Omicron wave when BA.5 became the predominant variant by August 2022 [2, 3]. In our analyses, severity estimates were compared between different epidemic wave periods based on the date of confirmation (Table S1). We also classified the period from 23 May 2022 onwards as wave 6 instead of wave 5 upon the predominance of the Omicron BA.4/BA.5 variants in late 2022.

Table S1. Classification of six COVID-19 epidemic waves in Hong Kong.

| Epidemic wave | Period | Predominant variant(s) |
|---|---|---|
| 1 | 23/1/2020 - 29/2/2020 | Ancestral |
| 2 | 1/3/2020 - 31/5/2020 | Ancestral |
| 3 | 1/6/2020 - 31/10/2020 | Ancestral |
| 4 | 1/11/2020 - 30/12/2021 | Ancestral |
| 5a (pre-peak period of wave 5) | 31/12/2021 - 13/2/2022 | Omicron BA.2 |
| 5b (peak period of wave 5) | 14/2/2022 – 29/4/2022 | Omicron BA.2 |
| 5c (post-peak period of wave 5) | 30/4/2022 – 22/5/2022 | Omicron BA.2 |
| 6a | 23/5/2022 – 30/8/2022 | Omicron BA.2 |
| 6b | 31/8/2022 – 26/10/2022 | Omicron BA.4/BA.5 |

### 2. Identification practices of confirmed cases in Hong Kong

In Hong Kong, all COVID-19 cases during the first four waves were confirmed through RT-PCR tests and hospitalized regardless of medical needs and severity of infection. Cycle threshold ($C_t$) values were initially used as part of the discharge criteria for hospitalized COVID-19 patients, but subsequently removed during the exponential growth phase between mid and late February 2022 (Table S2). From 20 February 2022 onwards, the decision to discharge hospitalized COVID-19 patients were subjected to the professional judgement of medical practitioners, and rapid antigen tests (RATs) were used to facilitate the discharge of patients who completed 14 days of isolation at community isolation facilities (CIFs) or their own homes. The case ascertainment was largely complete from wave 1 to wave 5a.

Table S2. Hospital-related policies throughout the COVID-19 pandemic.

| Date (wave) | Policy | Reference |
|---|---|---|
| 6/8/2021 (Wave 4) | Hospitalized patients are discharged if multiple clinical specimens taken at least 24 hours apart were tested negative via RT-PCR and test results show a $C_t$ value of at least 33. | [4] |
| 15/2/2022 (Wave 5b) | The Hospital Authority will prioritize hospitalization of preliminarily positive or confirmed patients with more severe symptoms or at higher risk. Meanwhile, patients waiting for hospital admission (typically mild or asymptomatic patients) are advised to self-isolate at home and await for further instructions. | [5] |
| 20/2/2022 (Wave 5b) | Whether a hospitalized patient can be discharged is up to the professional judgement of medical practitioners. Those who were discharged earlier than day 14 will be asked to stay home and can resume normal activities if test result on day 14 returns negative. | [6] |

**3. Data used in our study**

We only included local COVID-19 cases confirmed through either RT-PCR or rapid antigen tests (RAT) in our analyses since our study explored local transmission during the COVID-19 pandemic. We retrospectively classified the severity of all local COVID-19 cases based on available medical and epidemiological information from the datasets (Table S3). To ensure a comparable denominator for hospitalized cases despite changes in discharge or admission criteria (Table S2), we defined hospitalized cases as those classified with severity of severe or above and with confirmation-to-admission delays of no more than 14 days (Table S4). In addition to our main analyses, we compared the HFR estimates among hospitalized cases that did not receive any dose of the COVID-19 vaccine across different epidemic wave periods excluding the peak period of wave 5 (Table S1, Figure S1).

Table S3. Definitions for each severity classification and the number of local cases confirmed between waves 1-6 that meet the criteria.

| **Classification** | **Criteria** | **Waves 1-4** | **Wave 5a** | **Wave 5b** | | **Wave 5c** | | **Wave 6a** | | **Wave 6b** | |
|---|---|---|---|---|---|---|---|---|---|---|---|
| | | | | **PCR** | **RAT** | **PCR** | **RAT** | **PCR** | **RAT** | **PCR** | **RAT** |
| Total | - | 9,405 | 9,972 | 734,451 | 445,424 | 2,400 | 3,528 | 108,861 | 195,512 | 92,263 | 253,498 |
| Fatal | Encountered in-hospital fatalities | 208 (2.2%) | 48 (0.5%) | 7,898 (1.1%) | 547 (0.1%) | 42 (1.8%) | 5 (0.1%) | 467 (0.4%) | 68 (0.03%) | 398 (0.4%) | 100 (0.04%) |
| Critical | Ever required intubation or extracorporeal membrane oxygenation (ECMO) or in shock;<br>Ever admitted to ICU | 379 (4.0%) | 21 (0.2%) | 1,246 (0.2%) | 120 (0.03%) | 24 (1.0%) | 0 (0.0%) | 249 (0.2%) | 56 (0.03%) | 241 (0.3%) | 76 (0.03%) |
| Severe[a] | On treatment with Dexamethasone (+Remdesivir), and/or with either baricitinib or IV tocilizumab[b];<br>Ever being $O_2$ desaturated ($O_2$ desaturation level $\leq$ 90%);<br>Ever required oxygen supplement of 3 Litres per minute or more | 1,398 (14.9%) | 217 (2.2%) | 11,628 (1.6%) | 1,457 (0.3%) | 133 (5.5%) | 24 (0.7%) | 2,163 (2.0%) | 657 (0.3%) | 2,342 (2.5%) | 838 (0.3%) |
| Moderate[c] | With lowest $O_2$ desaturation level between 90-94%;<br>Ever prescribed with Remdesivir (without Dexamethasone), and/or Interferon beta-1b[b];<br>Ever visited A&E, with triage category being at least semi-urgent;<br>Ever prescribed with Paxlovid or Molnupiravir[d], regardless of being admitted to hospital or not | 3,670 (39.0%) | 2,020 (20.3%) | 21,106 (2.9%) | 9,405 (2.1%) | 397 (16.5%) | 238 (6.7%) | 14,552 (13.4%) | 17,842 (9.1%) | 14,914 (16.2%) | 29,864 (11.8%) |
| Mild | Not being classified as either fatal, critical or severe and without any specific treatment or evidence of $O_2$ desaturation, regardless of being admitted to hospital or not. | 3,750 (39.9%) | 7,666 (76.9%) | 692,573 (94.3%) | 433,895 (97.4%) | 1,804 (75.2%) | 3,261 (92.4%) | 91,430 (84.0%) | 176,889 (90.5%) | 74,368 (80.6%) | 222,620 (87.8%) |

[a] Severe cases referred to cases who met any of the listed criteria and also not meeting the criteria of severer (i.e. critical or fatal) cases.

[b] This is based on the local guideline for treating COVID-19 cases in Hong Kong.
[c] Moderate cases referred to cases who met any of the listed criteria and also not meeting the criteria of severer (i.e. severe, critical or fatal) cases.
[d] These two antiviral drugs started to be available in Hong Kong since February 25, 2022, with the primary purpose being provided to non-severe COVID-19 patients with highest risk of hospitalization (therefore categorized as “moderate” if not meeting other criteria of severer cases).

Table S4. Characteristics of local COVID-19 cases hospitalized in Hong Kong by epidemic waves.

| | **Waves 1-4**[a] (N=1,979) | | **Wave 5a**[a] (N=284) | | **Wave 5b**[a] (N=22,295) | | **Wave 5c**[a] (N=226) | | **Wave 6a**[a] (N=3,562) | | **Wave 6b**[a] (N=3,876) | |
|---|---|---|---|---|---|---|---|---|---|---|---|---|
| | No | % | No | % | No | % | No | % | No | % | No | % |
| Age, years | | | | | | | | | | | | |
| <18 | 29/1,979 | (1.5) | 18/284 | (6.3) | 491/22,295 | (2.2) | 8/226 | (3.5) | 242/3,562 | (6.8) | 227/3,876 | (5.9) |
| 18-44 | 243/1,979 | (12.3) | 30/284 | (10.6) | 646/22,295 | (2.9) | 17/226 | (7.5) | 205/3,562 | (5.8) | 226/3,876 | (5.8) |
| 45-64 | 782/1,979 | (39.5) | 64/284 | (22.5) | 2,504/22,295 | (11.2) | 26/226 | (11.5) | 539/3,562 | (15.1) | 548/3,876 | (14.1) |
| 65-79 | 652/1,979 | (32.9) | 87/284 | (30.6) | 5,986/22,295 | (26.8) | 74/226 | (32.7) | 1,001/3,562 | (28.1) | 1,216/3,876 | (31.4) |
| ≥80 | 273/1,979 | (13.8) | 85/284 | (29.9) | 12,667/22,295 | (56.8) | 101/226 | (44.7) | 1,575/3,562 | (44.2) | 1,659/3,876 | (42.8) |
| Unknown | 0/1,979 | (0.0) | 0/284 | (0.0) | 1/22,295 | (0.004) | 0/226 | (0.0) | 0/3,562 | (0.0) | 0/3,876 | (0.0) |
| Male | 1,122/1,979 | (56.7) | 155/284 | (54.6) | 12,671/22,295 | (56.8) | 121/226 | (53.5) | 1,985/3,562 | (55.7) | 2,146/3,876 | (55.4) |
| $O_2$ <95% among those with $O_2$ measured and documented | 818/954 | (85.7) | 180/198 | (90.9) | 11,224/12,741 | (88.1) | 179/192 | (93.2) | 2,359/2,581 | (91.4) | 2,586/2,820 | (91.7) |
| $O_2$ documented <95% among all hospitalized cases | 818/1,979 | (41.3) | 180/284 | (63.4) | 11,224/22,295 | (50.3) | 179/226 | (79.2) | 2,359/3,562 | (66.2) | 2,586/3,876 | (66.7) |
| Severity status[b] | | | | | | | | | | | | |
| Moderate | 1,251/1,979 | (63.2) | 192/267 | (71.9) | 9,759/20,755 | (47.0) | 136/200 | (68.0) | 2,451/3,431 | (71.4) | 2,733/3,733 | (73.2) |
| Serious | 296/1,979 | (15.0) | 18/267 | (6.7) | 1,939/20,755 | (9.3) | 11/200 | (5.5) | 286/3,431 | (8.3) | 328/3,733 | (8.8) |
| Critical | 230/1,979 | (11.6) | 10/267 | (3.7) | 703/20,755 | (3.4) | 6/200 | (3.0) | 165/3,431 | (4.8) | 182/3,733 | (4.9) |
| Fatal | 202/1,979 | (10.2) | 47/267 | (17.6) | 8,354/20,755 | (40.3) | 47/200 | (23.5) | 529/3,431 | (15.4) | 490/3,733 | (13.1) |
| Admission to discharge (Median) | 17 days | | 13 days | | 11 days | | 10 days | | 10 days | | 8 days | |
| Admission to death (Median) | 18 days | | 15 days | | 8 days | | 12 days | | 15 days | | 10.5 days | |

[a] Waves 1-4 (23/1/2020 – 30/12/2021); Wave 5a (31/12/2021 - 13/2/2022); Wave 5b (14/2/2022 – 29/4/2022); Wave 5c (30/4/2022 – 22/5/2022); Wave 6a (23/5/2022 – 30/8/2022); Wave 6b (31/8/2022 – 26/10/2022).

[b] Worst recorded status during hospitalization.

Figure S1: Age-specific estimates of the hospitalization fatality risk for COVID-19 among unvaccinated hospitalized local COVID-19 cases in Hong Kong by wave.

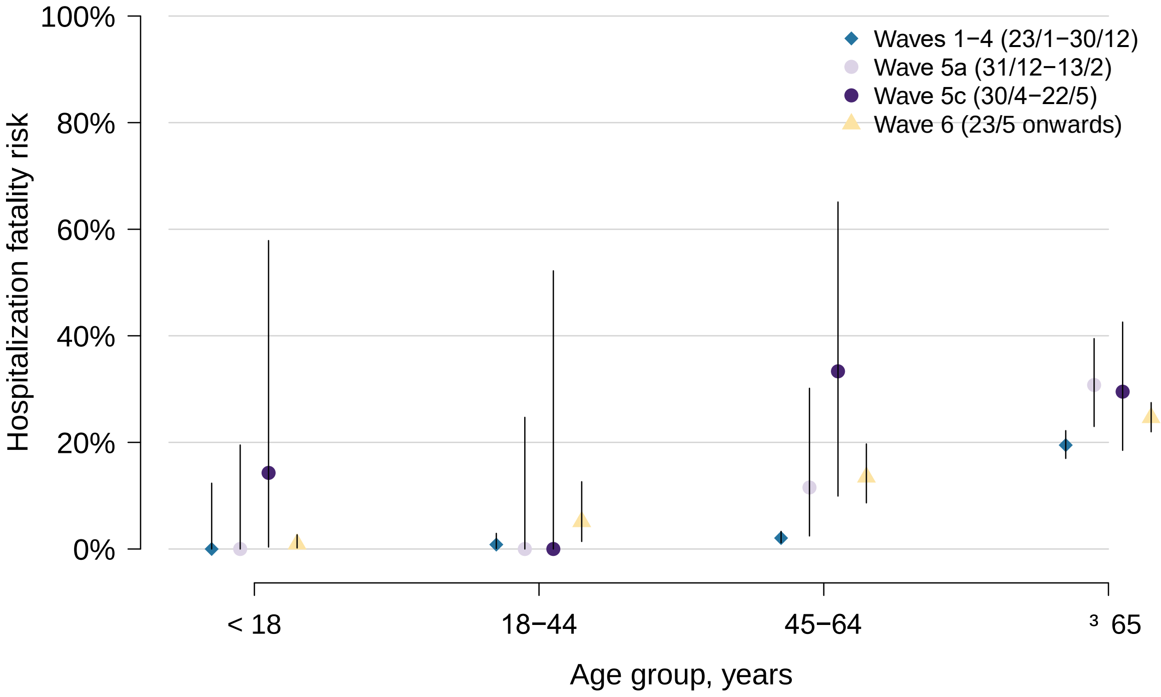

Figure S2: Fatality risk among hospitalized local COVID-19 cases in periods when Omicron BA.2 dominated (waves 5a, 5c and 6a) excluding the peak of the fifth wave, stratified by age group and vaccination status. (B) Relative risk of fatality among hospitalized cases with a specific vaccine dose combination compared to unvaccinated hospitalized cases by age group.

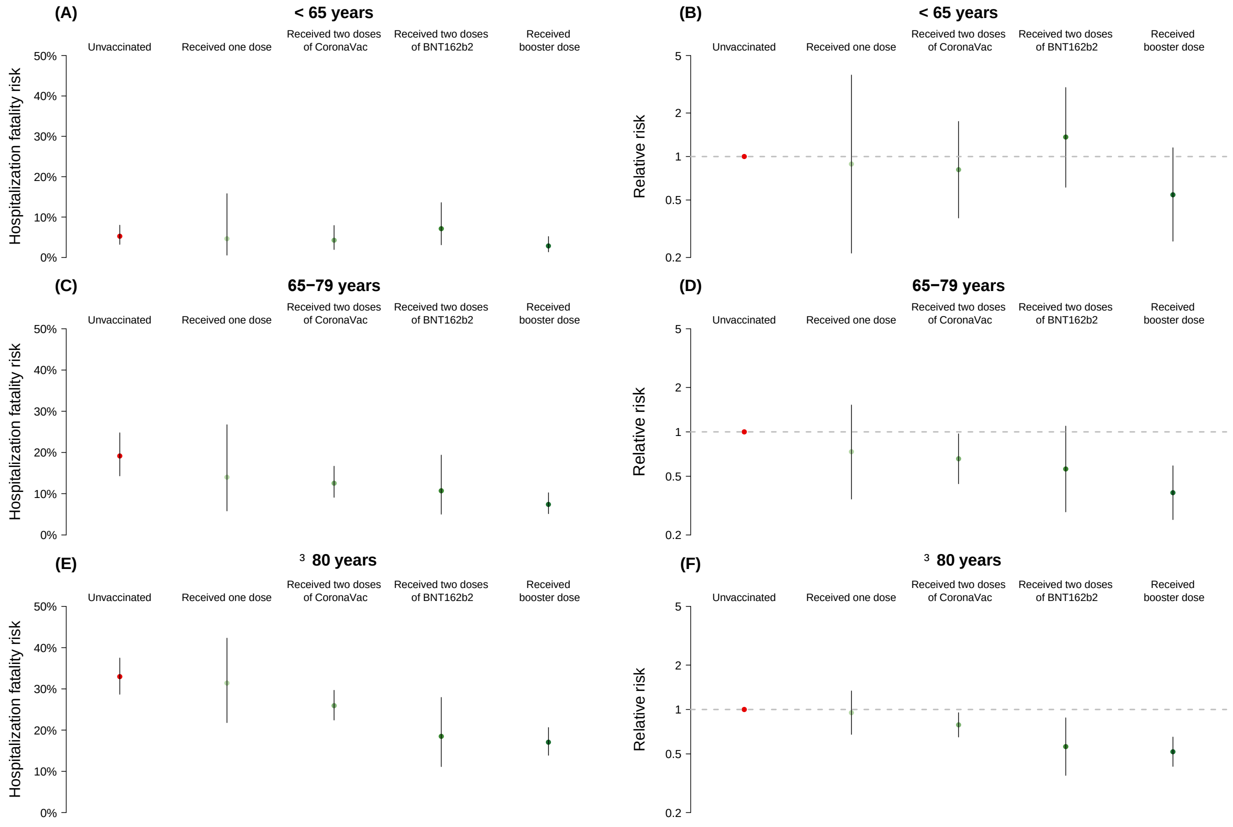
